# Black rims at 7 Tesla MRI: accumulation of iron around perivascular spaces in cerebral amyloid angiopathy

**DOI:** 10.64898/2026.04.22.26351134

**Authors:** Ivana Kancheva, Sabine Voigt, Leon Munting, Vera van Dis, Emma Koemans, Matthias J.P. van Osch, Marieke JH Wermer, Lydiane Hirschler, Marianne van Walderveen, Louise van der Weerd

**Author notes:** Joint first authors (Ivana Kancheva, Sabine Voigt). Joint senior authors (Marianne van Walderveen, Louise van der Weerd).

## Abstract

A prominent radiological manifestation of cerebral amyloid angiopathy (CAA) is enlargement of perivascular spaces (EPVS), which is suggested to result from fluid stagnation due to impaired perivascular clearance. Here, we report a novel observation of hypointense rims in cerebral white matter surrounding EPVS near haemorrhages on *in vivo* 7T Gradient Echo MRI. We hypothesised that the observed *‘black rim’* pattern denotes iron accumulation that may be caused by incomplete clearance following bleeding. We investigated the occurrence and localisation of this marker on *in vivo* and *ex vivo* MRI and examined its histopathological correlates.

From MRI data of the prospective longitudinal natural history study of hereditary Dutch-type CAA (D-CAA) at Leiden University Medical Centre, we selected the first 20 consecutive patients who underwent 7T imaging and assessed the presence of black rims on MRI. Post-mortem material was available from one donor with black rims on *in vivo* scans. Formalin-fixed coronal brain slabs were scanned at 7T MRI, including a high-resolution T2*-weighted sequence. Guided by *ex vivo* MRI, tissue blocks from representative areas with black rims were sampled for histopathological analysis. Serial sections were stained for iron, calcium, myelin, and general tissue morphology.

On *in vivo* 7T MRI, 9 out of 20 participants exhibited one or several black rims, all located close to a haemorrhage. In the D-CAA donor, *ex vivo* MRI signal loss matched the *in vivo* contrast changes. Thirty-six vessels with *ex vivo*-observed black rims were retrieved and histopathologically examined, showing iron accumulation surrounding perivascular spaces, but the pattern and severity of iron deposition varied. Across groups, vessels displayed microvascular degeneration, including hyaline vessel wall thickening, adventitial fibrosis, and perivascular inflammation.

We identified black rims on *in vivo* 7T MRI and confirmed their correspondence on *ex vivo* imaging. Iron deposition was determined as the underlying correlate of black rims, but the histopathology appears heterogeneous. The preferential deposition of iron around EPVS may indicate incomplete clearance of iron-positive blood-breakdown products after bleeding. The varied pattern of iron accumulation and microvascular alterations may reflect different pathophysiological mechanisms related to the formation and maintenance of black rims in D-CAA.

## Introduction

Cerebral amyloid angiopathy (CAA) is a major cause of lobar intracerebral haemorrhage (ICH) and cognitive decline in the elderly. It is characterised by a progressive deposition of amyloid beta (Aβ) in the walls of cortical and leptomeningeal arteries, likely due to impaired clearance of the peptide.^1,2^ CAA can occur sporadically in the general population as a subtype of small vessel disease, but it can also result from mutations in the amyloid precursor protein (APP) gene, one of which causes the autosomal dominant hereditary disorder of Dutch-type CAA (D-CAA, formerly known as hereditary cerebral haemorrhage with amyloidosis — Dutch type [HCHWA-D]).^3^ D-CAA and sporadic CAA (sCAA) are thought to be mechanistically comparable due to clinical, radiological, and pathological similarities between these forms of CAA.^4,5^

A definite diagnosis of CAA can only be made via post-mortem examination of brain tissue, but the Boston criteria aid CAA diagnosis during life, based on clinical symptoms and radiological findings.^6-8^ Imaging manifestations of sCAA and D-CAA may include haemorrhagic lesions on CT or MRI, including lobar and cortical microbleeds (CMBs), cortical superficial siderosis (cSS), and ICH^6,7,9,10^, as well as non-haemorrhagic injury of brain tissue, including lacunes, microinfarcts, and white matter hyperintensities (WMH).^11,12^ Additional phenomena have been detected using ultra-high-field 7 Tesla (7T) MRI, e.g., striped cortex and intragyral bleedings.^13^

A prominent radiological non-haemorrhagic manifestation of CAA is enlargement of the perivascular spaces (EPVS), commonly assessed in the centrum semiovale (CSO-PVS), which are compartments filled with interstitial fluid (ISF) surrounding small penetrating vessels. Increasing evidence suggests that PVS (also known as Virchow-Robin spaces) are involved in the exchange of solutes and removal of waste from the brain as part of a brain-wide cerebrospinal-fluid (CSF)-mediated brain clearance system.^14-16^ Generally, it is proposed that waste is transported from the ISF into the PVS of vessels before flow or mixing would drive it into the subarachnoid space where further elimination out of the cranium is facilitated.^17^ Other routes involved in compound exchange and clearance of solutes from the interstitium include the blood-brain-barrier (BBB) using active transporter mechanisms^18^ and perivascular macrophages.^19^ Although the exact pathways along which this process occurs remain unclear, in CAA, failure of ISF drainage and dilation of PVS have been proposed to result from impaired perivascular clearance of waste products such as Aβ, in cortical and leptomeningeal arteries. ^4,20,21^ Consistent with brain clearance dysfunction, previous studies have related CSO-PVS to lower amyloid levels in spinal CSF in D-CAA mutation carriers^22^ and to whole-cortex cerebrovascular amyloid burden in individuals with probable CAA.^23,24^

In this study, we report a novel imaging phenomenon originally identified on 7T MRI: hypointense rims around enlarged PVS in proximity to haemorrhages. The observed contrast on MRI scans suggested that rims denote accumulation of blood-breakdown products, possibly due to incomplete clearance following bleeding. Based on this observation, we aimed to investigate the frequency of this MRI marker *in vivo* and characterise its localisation in relation to haemorrhages, evaluate its correspondence on *ex vivo* 7T imaging, and examine its underlying histopathological substrates.

## Materials and methods

### Study population

Data were collected from individuals partaking in a prospective longitudinal natural history study: the AURORA study, which includes pre-symptomatic and symptomatic mutation carriers with D-CAA. Participants were recruited between 2018 and 2022 via the outpatient clinic of Leiden University Medical Centre (LUMC) in The Netherlands. Inclusion criteria were as follows: age of ≥18 years and a genetically confirmed APP mutation. Symptomatic D-CAA was defined by a history of at least one symptomatic ICH. Demographic information was obtained during standardised annual study visits. This investigation included the first 20 (both pre-symptomatic and symptomatic) consecutive participants enrolled during the recruitment period who underwent 7T MRI.

All participants provided written informed consent. The AURORA study was conducted in accordance with the Helsinki protocol and was approved by the Medical Ethics Review Committee of the Leiden-The Hague-Delft region (METC-LDD) in the Netherlands, which granted ethical approval for the study (reference number: NL62670.058.17). All procedures involving human participants were also approved.

### Magnetic Resonance Imaging

#### In Vivo Imaging

Scans were acquired at the LUMC on a whole-body human 7T MRI system (Philips, Best, The Netherlands) with a quadrature transmit and 32-channel receive head coil (Nova Medical, Wilmington, MA, USA) according to a previously published protocol.^13^ The sequence regarded as relevant for the current study consisted of a 2D flow-compensated transverse T2*-weighted Gradient Echo (GRE) scan with the following parameters: repetition time (TR)/echo time (TE) 1851/25 ms, flip angle 60 degrees, slice thickness 1.0 mm with a 0.1 mm interslice gap, 92 slices (multiband factor 2) and coverage of 10 cm, 240 × 180 × 100 mm field-of-view (FOV), 1000 × 751 matrix size – resulting in an in-plane spatial resolution of 0.24 × 0.24 mm, scan duration of 10 minutes. All participants also underwent 3T MRI performed using a standard 32-channel head coil (Philips, Best, The Netherlands), the parameters and details of which can be consulted in Koemans *et al*.^25^

Black rims were defined as areas of low signal intensity (dark) around MRI-visible perivascular spaces (i.e., EPVS) on susceptibility-weighted (SWI) or T2*-weighted GRE MRI images at 7T. In all participants, the presence of rims was evaluated by two independent observers (SV, >7 years of experience; LH, >7 years of experience). For descriptive purposes, MRI markers of cerebral small vessel disease were recorded at baseline 3T MRI according to the STRIVE criteria, including WMH, EPVS, cSS, and cerebral microbleeds.^26^

#### Brain Tissue

One of the patients who participated in the D-CAA natural history study deceased and generously donated brain tissue to the CAA brain bank maintained by LUMC. Written informed consent was acquired for the donor by their legal representative prior to autopsy. All material and data were handled in a coded manner maintaining patient anonymity according to the Dutch National ethical guidelines (Code for Proper Secondary Use of Human Tissue, Dutch Federation of Medical Scientific Societies). Figure 1 provides an overview of the retrospective post-mortem analytical workflow.

**Figure 1.**
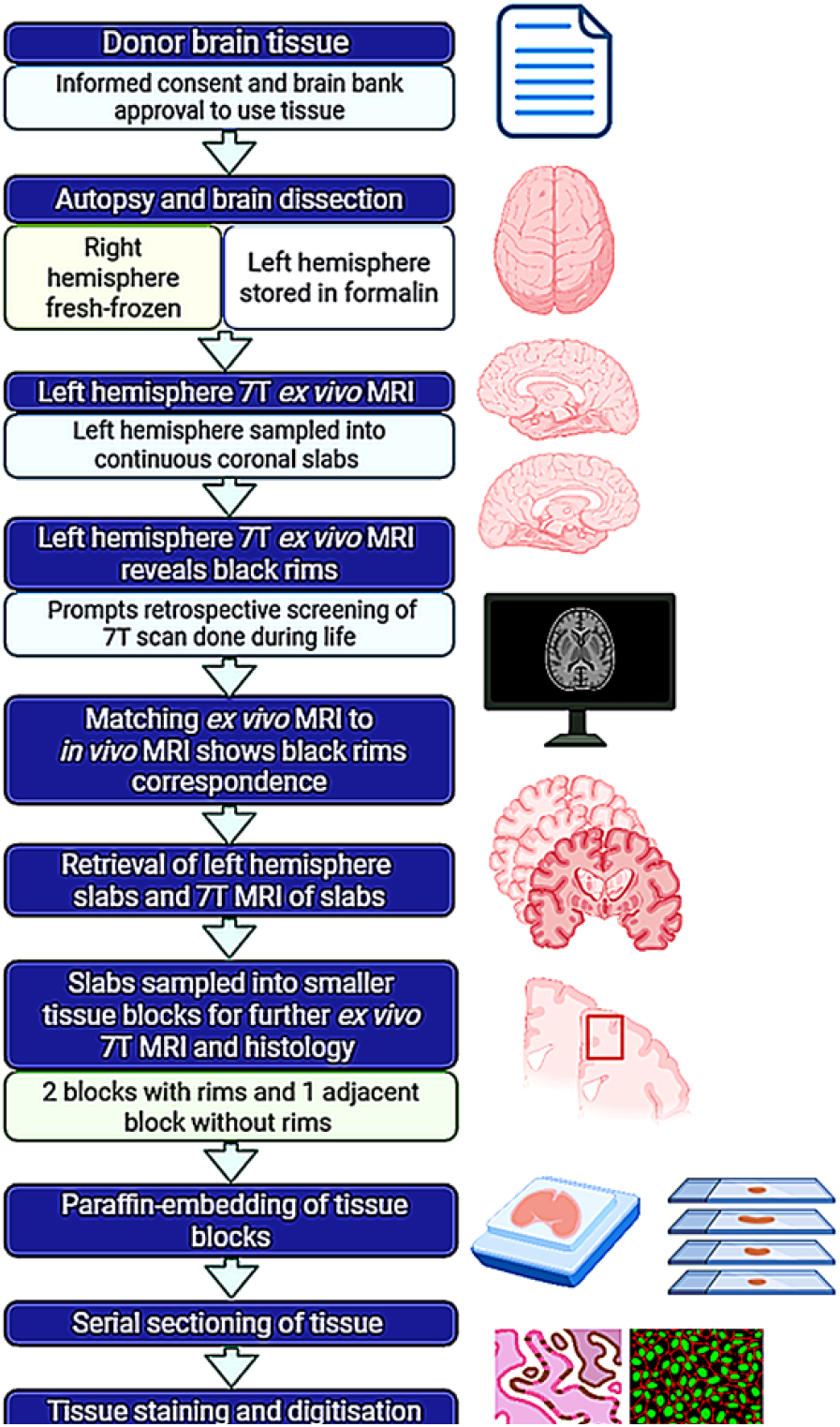
Overview of the post-mortem study workflow. A patient with D-CAA donated brain tissue to the LUMC brain bank after death. Approval was granted to use the tissue from the donor’s legal representative and CAA brain bank. At autopsy, the brain had been extracted, with material from the right hemisphere fresh-frozen and the left hemisphere fixed in formalin. The left hemisphere was subsequently scanned using high-resolution *ex vivo* 7T T2*-weighted Gradient Echo MRI and sectioned into 1-cm-thick continuous coronal slabs, which were stored in formalin. The *ex vivo* MRI revealed the presence of black rims, prompting a retrospective evaluation of the corresponding *in vivo* scan acquired during life, where matching signal loss was confirmed. After that, the previously stored slabs from the left hemisphere were retrieved and imaged at 7T, which guided subsequent definition of smaller tissue blocks (≈20 × 15 × 15 mm) containing rims, along with an adjacent control block without rims, that also underwent further *ex vivo* imaging before histopathological processing. The tissue blocks were paraffin-embedded, serially sectioned and stained using histological and immunofluorescent protocols, followed by digitisation. Finally, the stained and digitised sections were matched to *ex vivo* MRI through qualitative assessment and prepared for (semi)quantitative analysis. Figure created in https://BioRender.com.

#### Brain Tissue Preparation

At autopsy, the brain of the donor was extracted, and material was sampled from the right hemisphere for fresh-frozen tissue and fixed in 10 % formalin for at least 6 weeks. After that, the hemispheres were separated by a single midsagittal cut, and the left hemisphere was scanned *ex vivo*. Prior to scanning, the hemisphere was washed with phosphate buffered saline (PBS) for 24 hours to partially restore relaxation parameters. Afterwards, the tissue was placed in a plastic bag containing a proton-free fluid (Fomblin, LC08, Solvay), which was vacuum-sealed to eliminate trapped air bubbles. The plastic bag with the hemisphere was positioned on a custom-built plexiglass plateau that fitted in the centre of the head coil with the frontal lobe placed in the head direction and the occipital lobe in the feet direction of the scanner. Padding was used to prevent motion. The *ex vivo* scan was acquired overnight with the same human 7T MRI system used for *in vivo* imaging. Amongst others, the protocol included an overnight T2*-weighted GRE scan obtained with the following parameters: TR/TE 3140/25 ms, acquisition resolution 0.24 × 0.24 × 1 mm. After the scan, the left hemisphere was sampled into 1-cm thick continuous coronal slabs, which were arranged in the correct anatomical order and preserved in formalin. More details about the *ex vivo* post-mortem MRI protocols are described in Jonkman *et al*.^27^

The post-mortem scan of the left hemisphere revealed presence of black rims close to a macrohaemorrhage. This prompted inspection of the *in vivo* 7T MRI scan performed on the same patient during life.

#### Matching Ex vivo MRI with In Vivo Imaging

Retrospective evaluation of the *in vivo* MRI dataset acquired from the donor indicated the presence of black rims. Matching between *in vivo* and *ex vivo* MRI was carried out via visual inspection of the T2*-weighted GRE scans by one researcher (I.K.K.) whose assessment was cross-checked by three experienced observers (S.V., L.H., L.v.d.W.). Following confirmation of corresponding black rims on *ex vivo* and *in vivo* MRI, the previously sampled left hemisphere slabs that had already been stored were collected again for targeted *ex vivo* imaging and detailed histopathological analysis.

#### Post-mortem Image Acquisition

Two years after the hemisphere *ex vivo* scan, coronal brain slabs were scanned using the same procedure and protocol followed for the entire hemisphere. Subsequently, the *ex vivo* scans of the slabs were examined for the presence of rims by three independent observers (I.K.K., S.V., and L.H.), which guided the further resection of tissue into smaller blocks of ≈20 × 15 × 15 mm by a board-certified neuropathologist (V.v.D.). Three blocks were sampled to undergo additional *ex vivo* imaging. Two blocks included areas with identified rims; one adjacent block comprised areas where vessels with enlarged perivascular spaces without rims were observed, to serve as a control region. Before MRI, formalin was rinsed from the tissue blocks by washing under running tap water; then the blocks were individually placed in regular 50 mL tubes (Greiner Bio-One) filled with proton-free fluid (Fluorinert), which helps to minimise susceptibility artifacts in *ex vivo* brain MRI scanning. Care was taken to avoid trapped air bubbles by gently shaking the tissue. The scanning was also performed at the LUMC, but on a 7T horizontal bore Pharmascan MRI system (Bruker, Germany) with a 38-mm transmit-receive volume coil. The relevant sequence was a T2*-weighted Multi-GRE scan with parameters as follows: TR/TE 150/4.214 ms, flip angle 25 degrees, 43 × 30 × 15 mm FOV, 0.100 × 0.100 × 0.100 mm resolution with 8 averages, scan duration ≈ 11 hours and 20 minutes.

#### Tissue Sampling and Histology

The scanned tissue blocks were rinsed under running tap water before being fitted into tissue cassettes of appropriate size and thickness (≈0.2 – 0.5 cm). All samples were dehydrated, embedded in paraffin, and cut into serial sections. Deparaffinisation in xylene and rehydration through graded series of ethanol concentrations was done. For the detection of iron, two different protocols were used. The first method was an in-house developed protocol, based on the DAB-enhanced iron staining protocol described by Meguro *et al*.^28^, referred to as the Leiden iron staining.^29^ Briefly, 20-μm thick sections were subjected to sequential incubation steps in 1% potassium ferrocyanide (80 minutes), methanol with NaN_3_ and H_2_O_2_ (100 minutes), followed by incubation in 3,3’-DAB-tetrahydrochloride (40 minutes), with washing steps in-between. The second method was the classic Perl’s Prussian Blue for which sections were incubated with a 1:1 mixture of 4 % hydrochloric acid and 4 % potassium ferrocyanide (20 minutes), and counterstained with Nuclear Fast Red solution (10 minutes). Adjacent 6-μm sections were stained with Haematoxylin & Eosin (H & E) to assess vessel morphology, Von Kossa to detect calcification, and Klüver-Barrera Luxol Fast Blue (LFB) to assess white matter (WM) tissue rarefaction. To test for intracellularly stored iron, immunofluorescence against light-chain ferritin (1:100, ab69090, Abcam, USA) was carried out on consecutive sections. To tag light-chain ferritin, Alexa Fluor™ 647 (1:500) secondary antibody was used. Negative controls were included and exhibited no immunopositivity. A detailed description of the staining procedures is provided in Supplementary Methodological Note 1 and Supplementary Figure 1.

#### Matching Ex vivo MRI with Histology

For the *ex vivo* MRI-histology comparison, the most similar MRI slice with respect to the given histology was selected based on its physical position within the tissue block, estimated by counting the number of sections taken starting from the block’s surface. At this approximate location, matching of individual rims between MRI and histology was based on visual comparison of clearly detectable anatomical landmarks, i.e., grey-white matter border, contours, vasculature, tears, etc., and identifiable perivascular spaces. The scanned tissue blocks were systematically evaluated using the 3D Volume Viewer in Image J^30^ and every scan was rotated until an MRI slice was found that matched most closely the corresponding Leiden (DAB-enhanced) iron-stained histological section. Attention was paid to ensure that every rim identified on histology and related perivascular space could be tracked to the same vessel on the corresponding *ex vivo* MRI slice.

#### Histopathological Image Analysis

All histopathological slides of the donor were digitised using a 3D Histech Pannoramic 250 whole-slide scanner with a 20x objective (3D Histech Ltd., Budapest, Hungary). The digital images were exported and qualitatively assessed using Case Viewer software (version 2.9.0 for Windows 10). For fluorescent imaging, a Zeiss Axio Scan.Z1 whole-slide scanner was used, with consistent white balance and exposure time settings across sections to ensure standardised image acquisition, and Zeiss Zen Lite software (version 3.12) was employed for image viewing.

Every stained section was independently evaluated by two researchers (I.K.K. and L.v.d.W.) and an experienced neuropathologist (V.v.D.), blinded to MRI findings. The presence of iron was assessed on the sections stained with the Leiden (DAB-enhanced) iron protocol and Perl’s Prussian Blue. A rim was regarded as present if a hypointense rim could be consistently identified around an enlarged perivascular space in proximity to a haemorrhage on the respective *ex vivo* MRI slice, including across increasing echoes, and if this rim could be matched to an area of increased staining intensity on the histological section stained using the Leiden iron protocol. This decision was predicated on the higher sensitivity of the DAB-enhanced technique, based on the Meguro iron staining method, for identifying (non-haem) iron-positive structures in paraffin-embedded tissue.^28,29^ The localisation of iron accumulation was assessed (i.e., whether iron was deposited inside or around PVS). After it was established that iron was distributed around PVS, the pattern of iron accumulation severity was classified as follows: absent (–) if no increase in staining intensity was observed around PVS, mild (+/–) if patchy and/or inhomogeneous increase in staining intensity surrounding PVS could be detected, intermediate (+) if moderate increase in staining intensity surrounding PVS in a rim-like pattern was identified, or strong (++) if pronounced increase in staining intensity around PVS in a rim-like pattern was present, in consensus between the raters. Iron distribution was also qualitatively rated on every Perl’s section using similar criteria: absent (–) if no iron-positive deposits were found, mild (+/–) if some focal iron-positive deposits (blue pigment) were visible around PVS, intermediate (+) if moderate presence of iron-positive deposits was observed surrounding PVS, and strong (++) in case of substantial iron accumulation around PVS in a rim-like pattern, in agreement between the raters. This rating was supported by reference examples (Supplementary Figures 2 and 3). In addition, the presence of calcium and white matter rarefaction surrounding each vessel was assessed (yes/no). To characterise the vessels associated with rims in greater detail, the researchers and experienced neuropathologist also rated secondary degenerative changes on the H & E sections in consultation with previously published approaches^31,32^, following reference examples (Supplementary Figure 4). A consensus meeting was carried out to obtain a final score for each MRI-histology matched vessel with black rim. The inter-rater agreement prior to consensus was calculated as the number of agreements divided by the total rated vessels (×100 %). Lastly, to confirm the specificity of iron accumulation as the histopathological correlate of black rims, a tissue block comprising vessels with EPVS, but no rims on *ex vivo* MRI, was also rated.

## Results

### Population Characteristics

The median age of the participants was 53.5 (range 28 – 71 years) and 10 (50%) were women. Eleven participants (55%) had a history of symptomatic ICH (Table 1). Eight (40%) participants had cSS and 15 (75%) had CMBs (mean count: 196). Baseline characteristics and radiological findings of the D-CAA cohort are provided in Table 1.

**Table 1.**
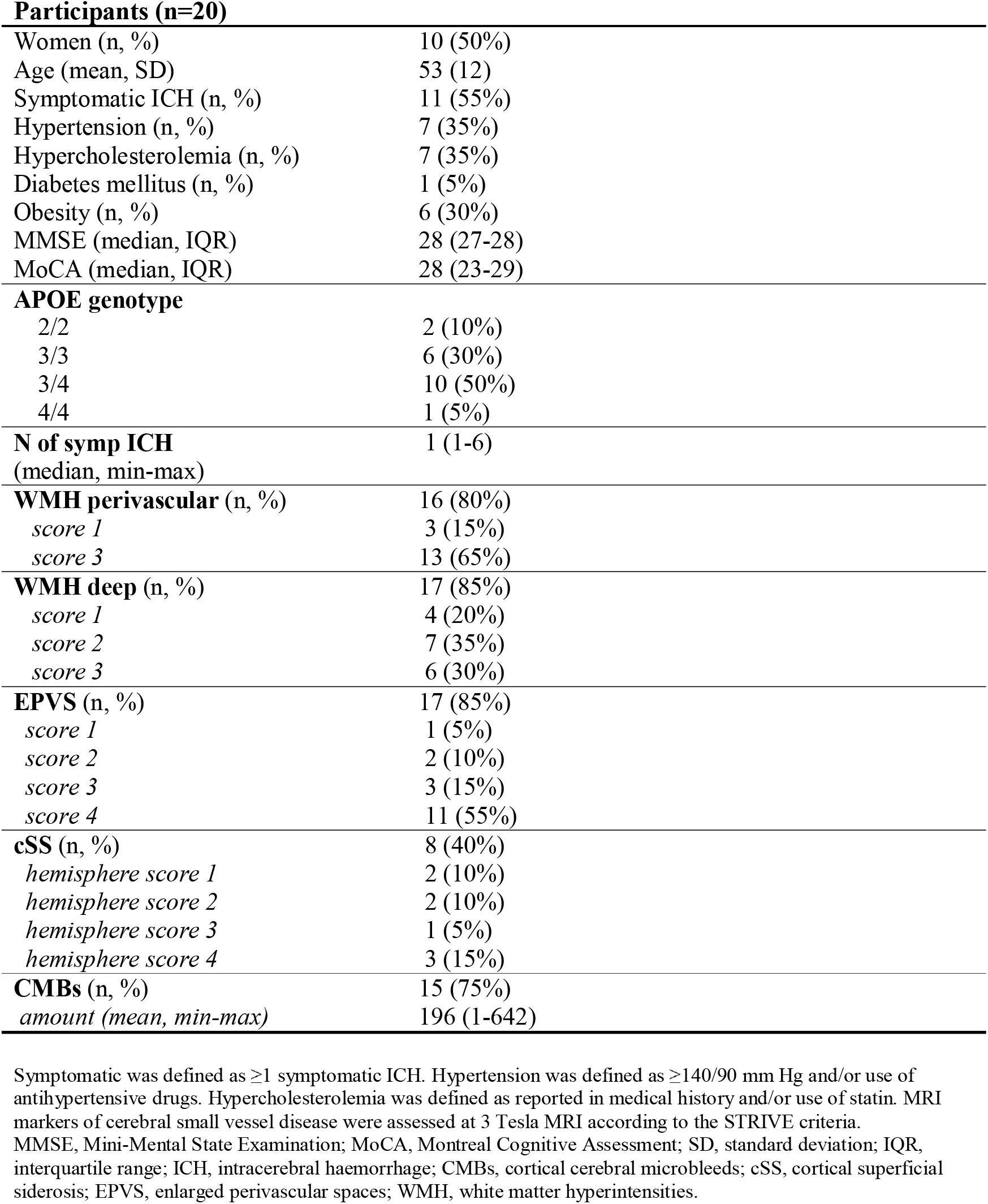
Baseline characteristics and radiological findings assessed on 3 Tesla MRI of D-CAA cohort.

### In vivo MRI Reveals Presence of Black Rims

In our cohort of 20 consecutive patients with D-CAA (Table 1), at least one black rim was found in nine participants (45%). In all cases, the rim was located next to a haemorrhage. When we subsequently inspected the 3T scans acquired on the same day, we could not distinguish the hypointense rims (Figure 2). In three of these nine participants, several macrohaemorrhages were present near the black rims and in one patient, cSS was observed near the rims. Seven patients had a symptomatic ICH. The total number of black rims was not quantified, as EPVS were not consistently visible across all regions and their detectability strongly depends on imaging orientation. However, whenever one or multiple black rims were observed, they were always in the vicinity of a bleed (mesobleed or larger macrohaemorrhage). Conversely, in regions with EPVS but no haemorrhages, no black rims were seen. In addition, we evaluated whether black rims could be distinguished in the absence of MRI-visible EPVS (i.e., without identifiable hyperintense CSF) on the *in vivo* 7T T2*-weighted GRE scans. No hypointense rims could be identified in areas without EPVS, however, this assessment is inherently challenging due to the hypointense appearance of vessels on iron-sensitive sequences and was further limited in most patients by extensive perihaematomal changes, presence of multiple haemorrhages, or cSS in proximal areas.

**Figure 2.**
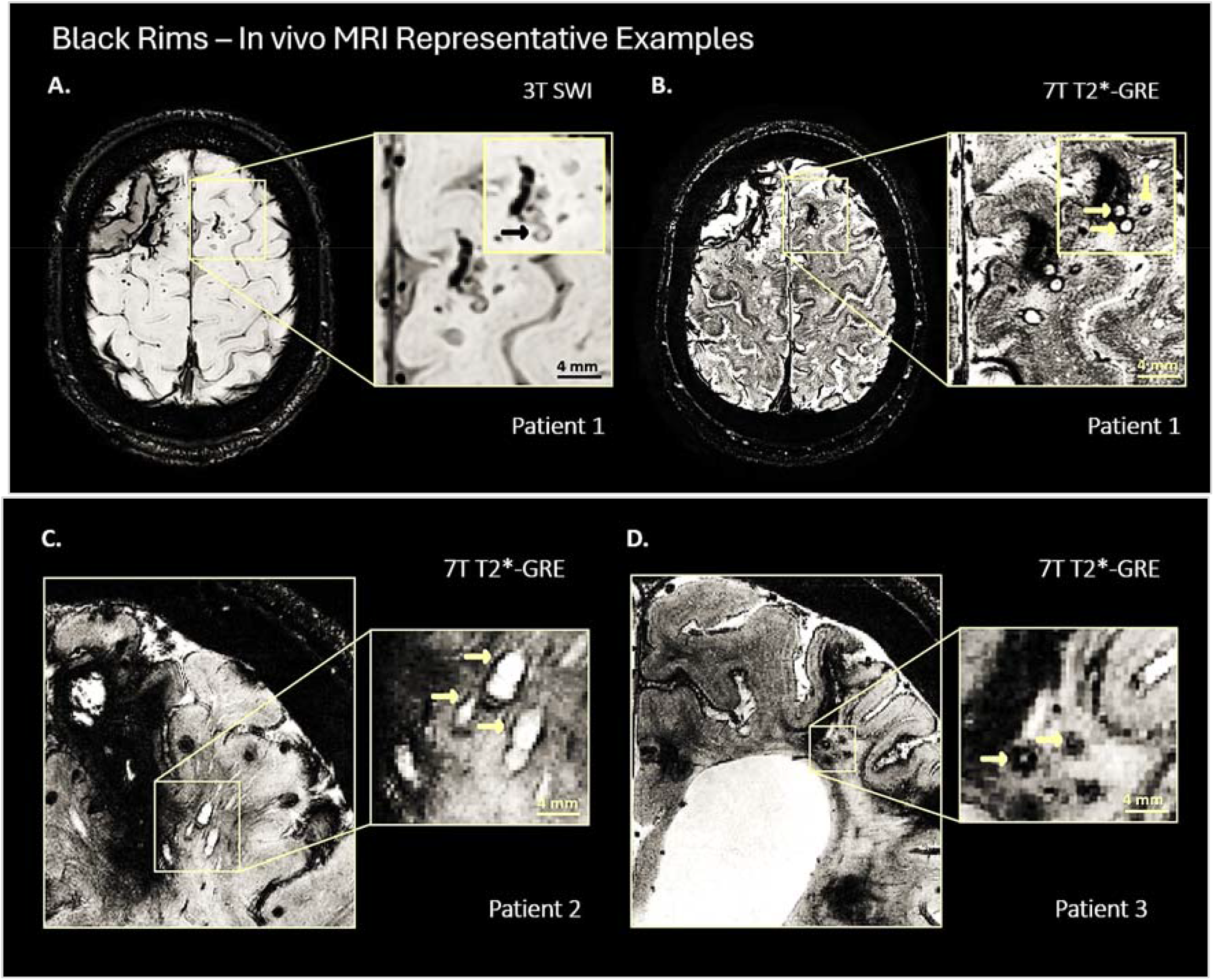
Black rims on In vivo iron-sensitive MRI. A representative area in a patient with symptomatic D-CAA shows a macrobleed and adjacent enlarged PVS in left frontal lobe (square in **A**). On *in vivo* susceptibility-weighted imaging (SWI) 3T MRI, no hypointense rims are visible, except subtle signal loss that could be identified around one vessel (**A**, *inset*, black arrow). The same area imaged using a high-resolution *in vivo* 2-dimensional transverse T2*-weighted Gradient Echo 7T MRI scan on the same day displays clearly discernible hypointense rims around the EPVS near the bleed (**B**, *inset*, yellow arrows). Panels **C** and **D** illustrate additional examples of black rims on 7T MRI in left occipital lobe in symptomatic D-CAA participants. The *insets* reveal greater level of detail. Scale bars indicate 4 mm.

### Correspondence of Black Rims on Ex Vivo and In Vivo MRI

One patient had deceased, and *ex vivo* 7T MRI data and post-mortem brain material were available for analysis in this study. The baseline and follow-up *in vivo* 7T T2*-weighted GRE scans of the patient are presented in Supplementary Figure 5. The interval between the last *in vivo* MRI and the whole-hemisphere *ex vivo* scan performed at autopsy was approximately 9 months. Autopsy confirmed D-CAA with extensive amyloid angiopathy (Thal phase 2 out of 3) without significant Alzheimer’s disease pathology, and no specific inclusions with alpha synuclein, ubiquitin, or tau.

On the *ex vivo* T2*-weighted GRE MRI scan of the left hemisphere, a black rim pattern was found around EPVS close to the haemorrhage in the targeted area of the frontal lobe, similar to the signal loss observed on *in vivo* MRI (Figure 3A and 3B). The same area was sampled into smaller tissue blocks for further *ex vivo* imaging, which confirmed the presence of black rims, and subsequent histopathological analysis (Figure 3C and 3D). On both *in vivo* and *ex vivo* MRI, no further rims were detected in other areas of this hemisphere.

**Figure 3.**
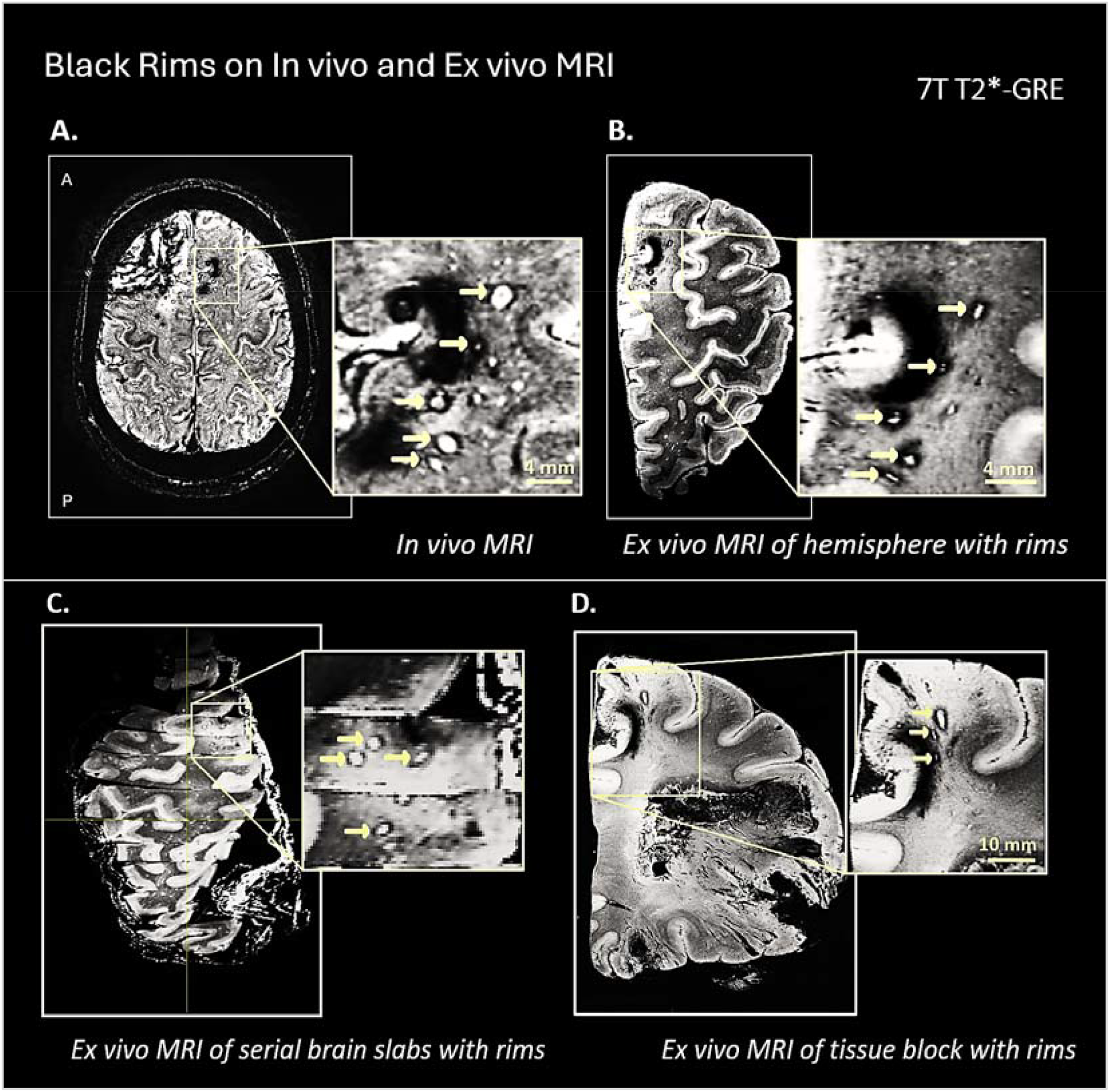
Black rims on In vivo and Ex vivo 7T iron-sensitive MRI. *In vivo* T2*-weighted Gradient Echo 7T MRI scan shows a bleed and adjacent enlarged PVS surrounded by hypointense rims in left frontal lobe in a patient with autopsy-confirmed D-CAA (**A**, *inset*, yellow arrows; axial view). *Ex vivo* 7T MRI of the same area in the left hemisphere reveals hypointense signal loss in a rim-like pattern, resembling the *in vivo* contrast changes (**B**, *inset*, yellow arrows). The scanned hemisphere is cut into continuous coronal slabs for further *ex vivo* imaging (**C**). The scanned slabs are resected into smaller tissue blocks, including targeted regions with rims, that undergo additional high-resolution scans (**D**), which confirm the presence of rims (yellow arrows in *insets*), and guide subsequent histopathological sampling. The *insets* reveal greater level of detail. A, anterior; P, posterior.

### Heterogeneous Histopathology of Black Rims

A total of 36 vessels from sampled areas with *ex vivo* MRI-visible rims were retrieved and matched to iron presence on histology, supporting iron accumulation as the underlying correlate of black rims (Figure 4). The inter-rater agreement for the (semi)quantitative assessment of these vessels before consensus was moderate-to-strong (77.80 %). Based on the severity of iron deposition and accompanying histopathological features, vessels with rims were classified into three groups.

**Figure 4.**
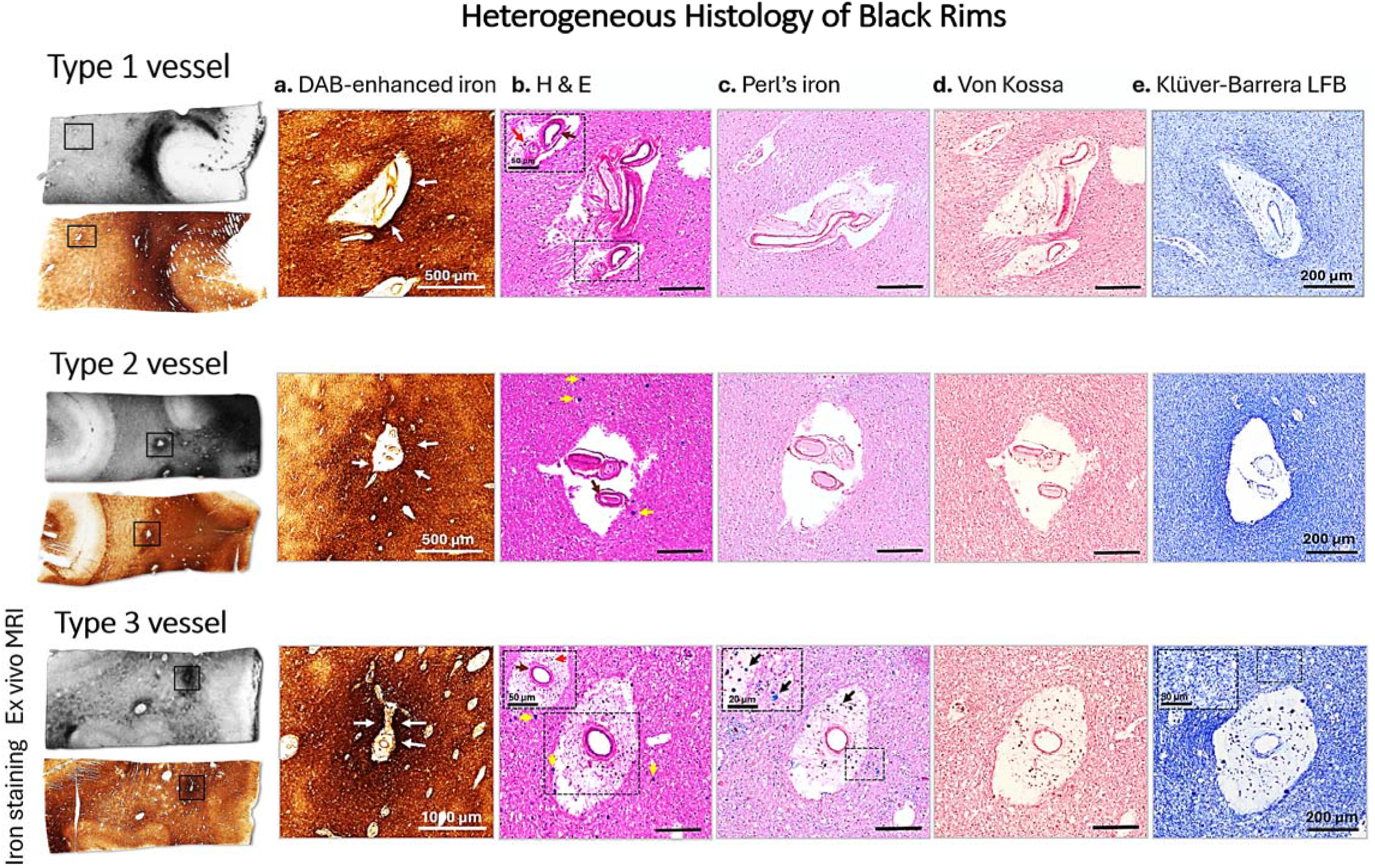
Heterogeneous histopathological correlates of black rims. Representative vessels with *ex vivo* MRI-visible black rims display heterogeneous patterns of iron accumulation on histology. **Top panel**. Type 1 vessel with a discrete hypointense rim on *ex vivo* MRI corresponds to a mild parenchymal iron deposition around PVS on Leiden (DAB-enhanced) iron staining (**A**, white arrows). Adjacent H & E (**B**) reveals intima thickening (*inset*, brown arrow) and immune cells with haemosiderin granules in a dilated perivascular space (*inset*, red arrow). Perl’s Prussian Blue iron (**C**) and Von Kossa calcium (**D**) stainings are negative. **Middle panel**. Type 2 vessel with a homogeneous hypointense rim on *ex vivo* MRI exhibits moderate iron accumulation around PVS on Leiden iron staining (**A**, white arrows). Vessel wall hyalinosis with marked PVS enlargement (**B**, brown arrow), infiltration of macrophages/histiocytes within the vessel wall, and *corpora amylacea* within 0.5 mm of the PVS (**B**, yellow arrows) are visible. Perl’s Prussian Blue staining (**C**) is negative, and no calcification (**D**) or WM rarefaction (**E**) are observed surrounding the vessel. **Bottom panel**. Type 3 vessel shows strong Leiden iron staining (**A**, white arrows) with severe microvascular degenerative changes (**B**). Haemosiderin deposits are visible on Perl’s around, and to a lesser extent, inside the PVS (**C**, *inset*, black arrows) without calcification (**D**). Asymmetrical WM rarefaction in the neuropil surrounding the vessel can be seen (*inset* in **E**), suggestive of WM injury. Panels A-E correspond to the areas indicated with squares on the MRI images. **Scale bars: A** (top and middle) = 500 µm; **A** (bottom) = 1000 µm; **B–E** = 200 µm. Insets show higher magnification. H & E, Haematoxylin and Eosin; LFB, Luxol Fast Blue.

Nine vessels were classified as Type 1 (Figure 4, top panel). These vessels exhibited variable signal loss on *ex vivo* MRI – either inhomogeneous or discrete hypointense rim. Seven of these vessels were matched to areas where mild increase in staining intensity was seen on the Leiden iron staining around, but not inside the PVS, and for two the latter appeared either mildly stained or inconclusive. No iron accumulation was seen on Perl’s, and no calcification was evident.

The second group comprised 21 vessels classified as Type 2 (Figure 4, middle panel). They were located close to the sites of bleeding identified on the matched *ex vivo* MRI slices, but not immediately proximal. All vessels showed intermediate or strong parenchymal iron surrounding the PVS on the Leiden iron protocol-stained sections and corresponded to clearly defined hypointense rims on *ex vivo* MRI. Perl’s was either negative or revealed minimal focal haemosiderin deposits. On the H & E staining, moderate microvascular degenerative and inflammatory changes were present in most cases (13 out of 21 vessels) and wasteosomes (*corpora amylacea*) were noted around several vessels. No calcification or WM tissue rarefaction were observed surrounding these vessels.

The third group included six vessels located near the macrohaemorrhages visible on the matched *ex vivo* MRI slices, which corresponded to intermediate or strong parenchymal iron around PVS on both the Leiden iron protocol and Perl’s. They were classified as Type 3 vessels (Figure 4, bottom panel). Four of them exhibited severe microangiopathic alterations on H & E and presence of *corpora amylacea* surrounding the PVS. White matter rarefaction around these vessels was not evident. The histopathological findings of all retrieved vessels with rims are summarised in Table 2. A more detailed description is available in Supplementary Table 1.

**Table 2.**
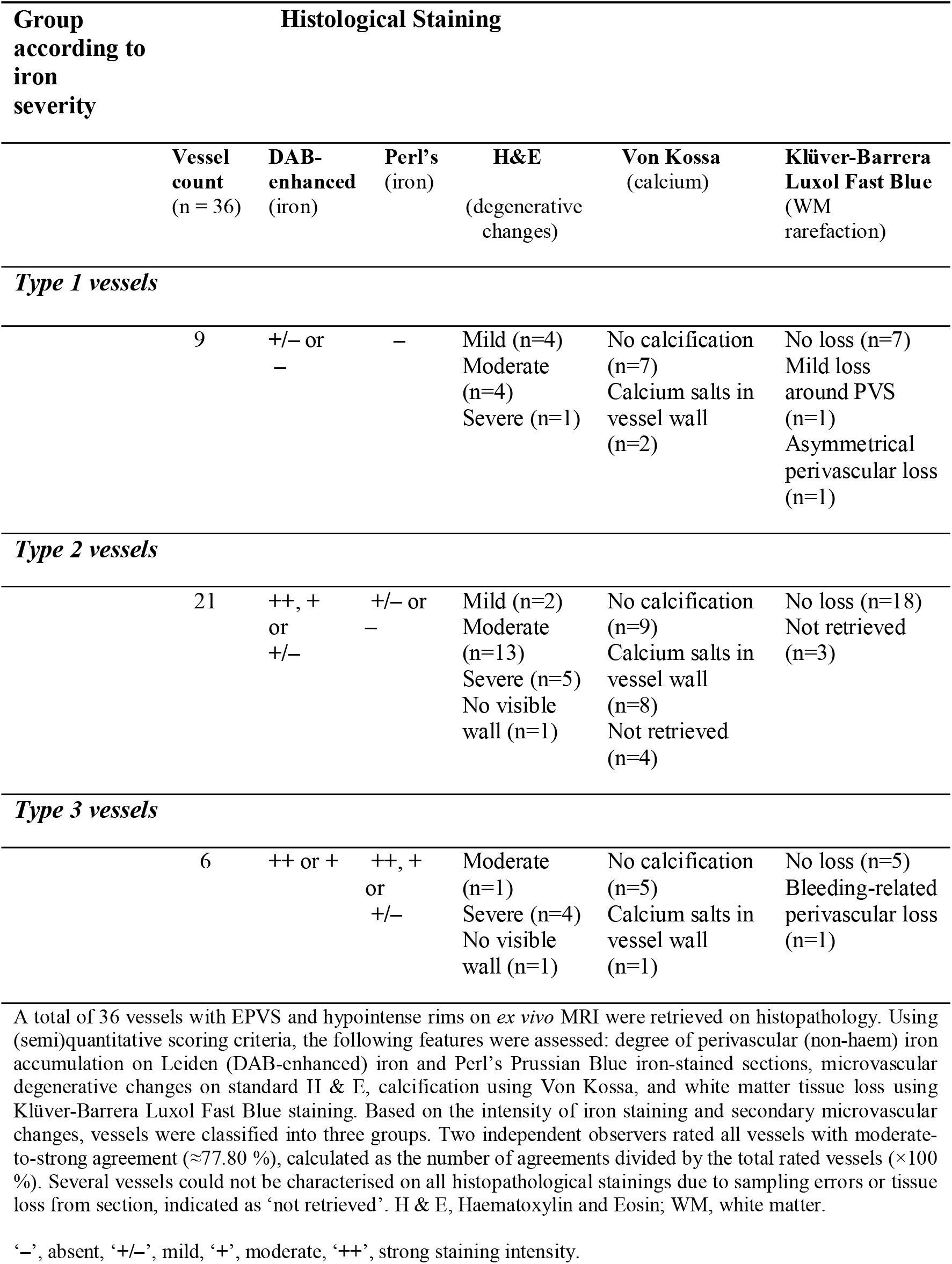
Histopathological correlates of *ex vivo* MRI-observed black rims.

In a subset of vessels with a positive Leiden iron staining, we additionally carried out immunofluorescence against light-chain ferritin, which revealed upregulation of iron-storage complexes in cells surrounding the rims, closely matching the iron distribution seen using the DAB-enhanced protocol (Figure 5).

**Figure 5.**
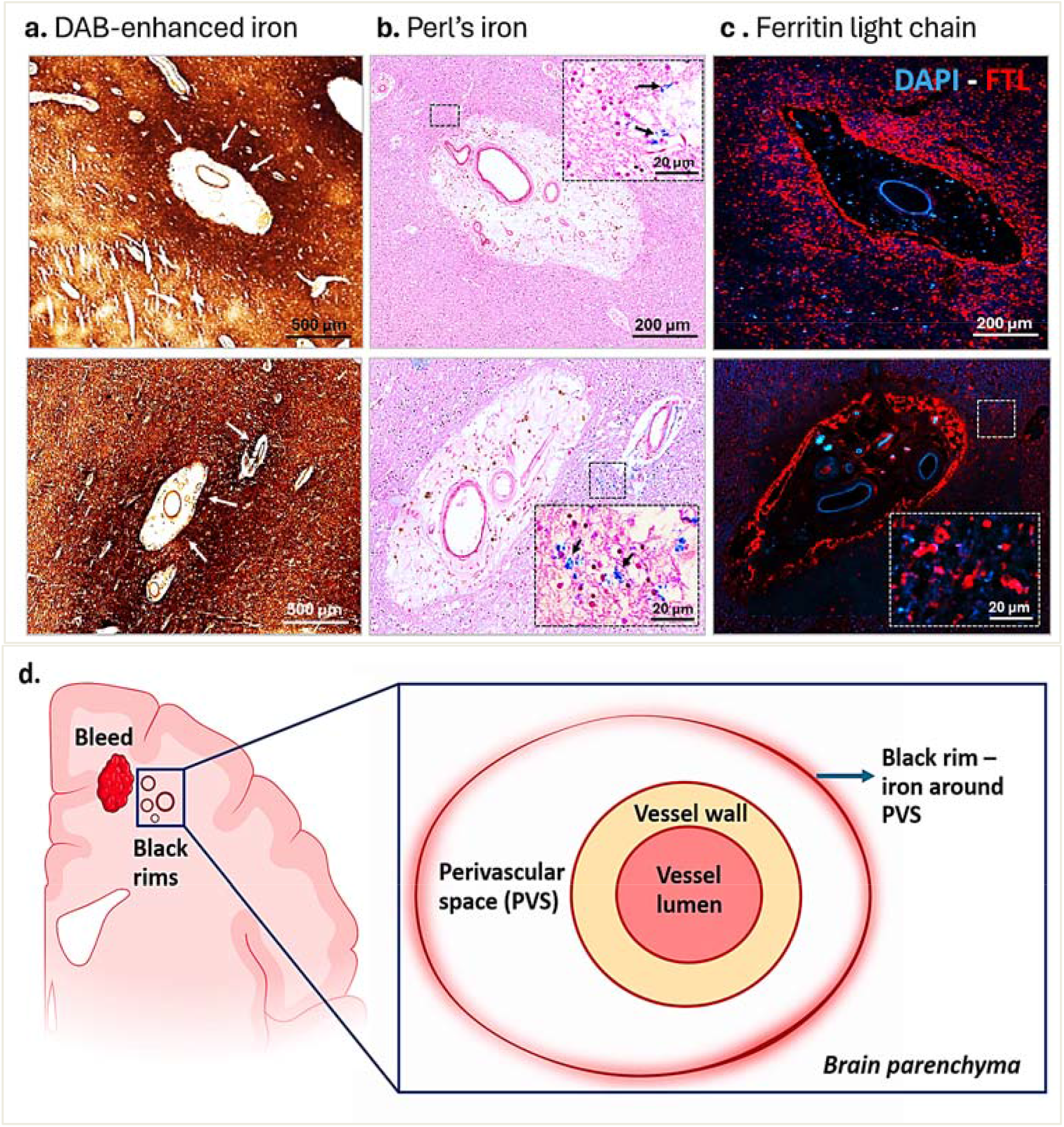
Distinct patterns of iron accumulation around PVS on histology. **Top panel**. Representative Type 2 vessel (top row) and Type 3 vessel (bottom row) exhibit strong iron staining intensity surrounding PVS on Leiden (DAB-enhanced) iron staining (**A**). Only patchy focal deposits of haemosiderin are visible on Perl’s (**B**, *insets*, black arrows). Immunofluorescence against light-chain ferritin (**C**) reveals multiple iron-positive inclusions around the PVS that closely match the pattern seen on the Leiden (DAB-enhanced) iron protocol. **Bottom panel**. Schematic of a vessel with black rim, formed by accumulation of iron around an enlarged perivascular space in cerebral white matter close to a bleed (**D**). The illustration represents a simplified conceptual interpretation of the parenchymal iron distribution around perivascular spaces observed in A. **Scale bars: A** = 500 µm; **B, C** = 200 µm. Insets indicate 20 μm magnification. FTL, ferritin light chain; PVS, perivascular space.

Furthermore, six vessels with mildly enlarged PVS but without hypointense rims on *ex vivo* MRI were sampled to serve as controls. No iron deposits were observed in the parenchymal areas surrounding these vessels (Supplementary Figure 6 and Supplementary Table 2). Lastly, all vessels with EPVS and black rims on *ex vivo* MRI could be retrieved on histopathology. However, several vessels where mild parenchymal iron surrounding PVS was detected on the Leiden iron staining could not be reliably matched to hypointense rims on *ex vivo* MRI due to their small size (Supplementary Figure 7). Additionally, two vessels located close to a haemorrhage displayed black rims on *ex vivo* MRI, corresponding to iron-positive blood products *inside* the PVS, rather than around it, on histology (Supplementary Figure 8). These vessels were not rated.

## Discussion

We report a new imaging observation on ultra-high-field iron-sensitive MRI, termed the *black rim*, in participants with D-CAA. We examined the frequency and localisation of this MRI phenomenon *in vivo*, its correspondence with *ex vivo* 7T imaging, and its underlying histopathology. Our combined approach revealed several key findings: in all participants with black rims identified *in vivo* around EPVS, these occurred close to haemorrhages; in one donor with available post-mortem material, *ex vivo* MRI matched the *in vivo* contrast changes; parenchymal iron accumulation around EPVS was confirmed as the neuropathological correlate of black rims, but the iron deposition patterns varied.

The consistent distribution of rims around EPVS and near haemorrhages may indicate several processes. One possibility is that rims represent localised accumulation of non-haem iron-positive blood-breakdown products, reflecting incomplete perivascular clearance following bleeding. Given that PVS are thought to constitute the principal route for clearance of interstitial fluid and solutes, including soluble amyloid β^33,34^, impaired function of these pathways may contribute to deposition of blood-derived material near haemorrhages. This is consistent with evidence relating PVS enlargement to impaired interstitial fluid drainage caused by leptomeningeal and cortical vascular amyloid β in sCAA^21,35,36^ and in ageing and AD models with CAA pathology.^37,38^ Previously, 7T iron-sensitive MRI has been used to detect, e.g., cortical microbleeds in CAA^39^ and cortical vascular iron accumulation and calcification of amyloid β-positive penetrating arteries in D-CAA.^13,40-42^ In line with these observations, rims were visible on 7T, but not 3T MRI *in vivo*, likely due to differences in sensitivity to superparamagnetic sources and spatial resolution. However, the iron deposition around EPVS in white matter described here has not been examined before and should be differentiated from the intravascular or cortical iron reported in these studies.

The underlying histopathology of rims was heterogeneous and implies distinct mechanisms related to blood clearance following haemorrhage. A subset of vessels (Type 1) exhibited subtle signal loss on *ex vivo* MRI, corresponding to mild parenchymal iron staining around PVS on the Leiden iron protocol, and negative Perl’s. These vessels may denote the presence of loosely bound or low-concentration non-haem iron in extracellular compartments, compatible with evidence that the DAB-enhanced (Meguro-based) method is more sensitive to extracellular and myelin-associated iron, whereas Perl’s Prussian Blue primarily stains ferric iron in haemosiderin and ferritin and may underestimate diffuse parenchymal iron.^28,29,40^

The largest group of retrieved vessels (Type 2) displayed clearly identifiable rims on *ex vivo* MRI. These were matched to areas of moderate-to-strong parenchymal iron located outside the PVS boundary. The distribution of iron around PVS supports the notion that paramagnetic substances from proximal macrobleeds spread along perivascular pathways and aggregate in haemorrhagic foci that appear as rims on iron-sensitive MRI. Prior investigations in ICH have demonstrated that haematoma contents can extend along compartments surrounding vessels and form perihaematomal haemorrhagic lesions. These extensions may be facilitated by the low-resistance perivascular spaces, which provide a preferential route for the spread of soluble toxic components, including iron.^43,44^. The absence of parenchymal membranes investing PVS in the WM and their enlargement may also enable the migration of haemorrhagic blood contents through the parenchyma, as previously suggested. ^34,45-47^ It is conceivable that our rims reflect a similar process where blood extravasation from adjacent bleeds expands along the PVS, but has difficulty diffusing further in the parenchyma, thus producing local inhomogeneous signal, visible as rims. A comparable mechanism could explain the histological appearance of Type 3 vessels where parenchymal iron around PVS was visible on both iron protocols we used. The greater intensity detected by Perl’s likely reflects higher concentrations of haemosiderin-rich blood degradation products, consistent with the closer proximity of Type 3 vessels to macrohaemorrhages. Interestingly, *corpora amylacea* were identified around these vessels, which are proposed to increase at sites with excessive waste accumulation when glymphatic function is compromised.^48,49^

In just two vessels, hypointense rims corresponded histologically to haemorrhagic material *inside* the PVS. Experimental studies have described entry of blood-derived components throughout subarachnoid and arteriolar perivascular spaces following SAH^50-52^ and microinfarct-induced BBB disruption^53^ that get cleared by perivascular and meningeal macrophages. Our cross-sectional observations do not allow assessment of whether iron has transited through the PVS before being taken up in the surrounding parenchyma to form rims. As such, they do not provide evidence for any specific brain clearance route but may be considered as hypothesis-generating with regards to mechanisms and directionality of perivascular clearance.

Another feature we observed was moderate-to-severe microvascular degeneration of vessels with black rims, which may relate to greater CAA severity in the overlying cortex, or denote secondary microangiopathic and inflammatory changes caused by presence of parenchymal blood-breakdown products. Further investigations are needed to clarify the link between microvascular and haemorrhagic injury, especially given the paucity of knowledge about vessel alterations in cerebral white matter in CAA.

A strength of this work is the integration of *in vivo* and *ex vivo* ultra-high-field MRI and detailed histopathology to characterise a novel imaging observation in CAA. One limitation is our population of a selective group of patients enrolled in D-CAA natural history studies at LUMC, which may not generalise to other settings or to sporadic CAA. Prospective longitudinal imaging, integrated with clinical outcome measures in larger cohorts, could elucidate processes that contribute to rim formation. Other limitations include the availability of tissue from a single donor and the cross-sectional nature of our observations that only capture static non-temporal information, which is an inherent drawback of post-mortem investigations. Future longitudinal studies combining *in vivo* MRI with time-resolved histological sampling in experimental models are needed to gain mechanistic understanding of perivascular clearance of blood components. Despite these drawbacks, it is worth acknowledging the rare availability of datasets with high-quality *in vivo* and *ex vivo* MRI and brain autopsy in D-CAA. Taken together, our findings highlight the importance of understanding clearance of blood products after haemorrhage. Future studies involving retrospective screening of iron-sensitive *in vivo* scans, detailed imaging, and demographic information from larger populations, including sCAA, may help disentangle the processes that lead to the formation and maintenance of black rims, and their significance for clinical CAA severity.

## Data Availability

Data that support the findings of this study are not publicly available to protect patient confidentiality and anonymity but can be obtained from the corresponding author upon reasonable request.

## Acknowledgements

The authors would like to thank Ingrid Hegeman, Ernst Suidgeest, and Kyra Dijkstra for excellent technical support. The authors wish to also thank all participants for their contribution to this research and to the family and legal representative(s) of the donor for generous brain donation.

## Notes

### Competing Interest Statement

The authors have declared no competing interest.

### Funding Statement

The AURORA study was funded by the Dutch Heart Foundation Dekker Grant (number: 2016T86). The project was also supported by several other research grants: the Dementia Research Programme (OPD) of ZonMw as part of the Mechanisms of Dementia (MODEM) consortium; Alzheimer Nederland grant (number: 77522) to LvdW; Alzheimer Nederland grant (number: WE.06-2023-07) to LPM; ENW VIDI grant (VI.Vidi.243.246, https://doi.org/10.61686/QCTYT86439) to LH. MJH Wermer receives support from the Dutch Heart Foundation (grant number: 2016T086). MJPvO and LH received support from a grant from the Leducq Foundation and the Leducq Foundation for Cardiovascular Research (number: 23CVD03) and from the project CHIME with file number P22.012 of the research programme Perspectief, which is (partly) funded by the Dutch Research Council (NWO).

### Author Declarations

The AURORA study was conducted in accordance with the Helsinki protocol and was approved by the Medical Ethics Review Committee of the Leiden-The Hague-Delft region (METC-LDD) in the Netherlands, which granted ethical approval for the study (reference number: NL62670.058.17).

